# Exploring Socio-demographic and Geographical Variation of Adults Hypertension in Bangladesh: Spatial Hotspot Analysis

**DOI:** 10.1101/2024.07.02.24309855

**Authors:** Anika Tabashsum, Chowdhury Mashrur Mahdee, Most. Jannatul Ferdous Asha, Md. Ashraful Islam Khan, Md. Aminur Rahman

## Abstract

Hypertension is a chronic medical condition where blood pressure is too high. In Bangladesh, the overall incidence of hypertension is rising, like in other developing countries. Recent studies show hypertension can cause other non-communicable diseases such as cardiovascular diseases, strokes, etc. Utilizing information from the Bangladesh Demographic and Health Survey (BDHS) 2017–18, a cross-sectional and spatial analysis was done to determine the prevalence of hypertension. The existence of spatial autocorrelation is evaluated by using Moran’s I statistic, hotspot analysis, cold spot analysis, and influential observation analysis. The study was conducted on the basis of different factors such as gender, place of residence, age, BMI, etc. Among 12,926 people (43.19% men, 56.81% women), the women had a higher total prevalence of hypertension (41.18% (95% CI, 41.35–45.67) compared to the men’s 58.82% (95% CI, 57.16–60.45). Out of the total number of observations, 27.47% had hypertension. Out of all hypertensive people, 57.56% are unaware of their condition, which is very concerning. The study also finds that 28.4% are from urban areas and 71.6% are from rural areas. People living in the Dhaka division had a higher prevalence of hypertension (20.5%), and those in Sylhet had the lowest prevalence of hypertension (6.2%). There also exists a statistically relevant spatial autocorrelation (hypertension: Moran’s I index = 0.27, p<0.001). These are the spatial clustering maps, and according to the LISA cluster map, the Northeast districts are the cold spots (8), while the hotspots (4) are Bagerhat, Rajbari, Pirojpur, and Munshiganj. Approximately more than one-fourth of the adult population of Bangladesh had hypertension. The analysis discovered the hotspots and cold spots of the prevalence of hypertension. These findings could facilitate awareness and treatment of hypertension and therefore serve as support for researchers and policymakers.

## Introduction

Elevated blood pressure, also known as hypertension, is a critical health problem that increases the chance of getting kidney, heart, brain, and other disorders.[1]. Additionally, elevated blood pressure can be used as a marker for other noncommunicable disease (NCD) risk factors, such as gaining weight, dyslipidemia, glucose intolerance, and the metabolic syndrome. 13% of deaths worldwide are mainly caused by hypertension[2]. Hypertension has emerged as one of the most challenging issues for global public health due to the prediction that the prevalence of the condition will rise by 30% by 2025, which is crucial given the increasing global disease burden and disability[3,4].

Worldwide, 1.13 billion people are hypertensive. According to recent trends, the prevalence of hypertension among people in low- and middle-income nations (LMICs) is rising at an alarming rate[5]. On the basis of studies, LMICs have a higher prevalence of hypertension (31.5%) than high-income countries (28.5%), with the majority of the world’s hypertensive population living in these regions[6]. There is no exception to this rising tendency in LMICs in South Asia, like Bangladesh[7–9]. The overall hypertension prevalence was first recorded for Bangladesh in 1976 at 1.10%[10]. According to a comprehensive study and meta-analysis of data covering the years 1995–2009, the hypertension prevalence in the country was 12.8% in men and 16.1% in women[11]. And roughly 28.4% of women and 26.2% of men who were at least 18 years of age or older have hypertension, depending on the latest Bangladesh Demographic and Health Survey (BDHS) 2017–18, which corresponds to the population of Bangladesh[9].

The ongoing nutritional shift from a standard diet to processed and fast food, rising trends of unhealthy lifestyles because of increased socioeconomic status, crowded living conditions, and a lack of physical activity as a result of unplanned urbanization may all play a significant role in the rise of the high blood pressure epidemic in Bangladesh[12]. Although major communicable diseases have been successfully eliminated in Bangladesh, the country’s health care system is facing a significant challenge from this new pattern of diseases.

In this country, the prevalence of hypertension is continuing to increase, and this needs to be controlled as soon as possible. Additionally, according to the BDHS report, over half of people with hypertension are unaware of their health condition[9]. So, it’s crucial to raise awareness of hypertension among individuals who have it and improve blood pressure (BP) management among people who use any kind of antihypertensive medication in order to lessen the burden of consequences emerging from it in the future[13]. Hence, it is needed to identify the causes or factors that contribute to the prevalence, awareness, and management of hypertension and to treat them through prevention programs. According to underlying characteristics and variables related to these outcomes in recent years, it is unknown what percentage of people have a prevalence, knowledge, or treatments for hypertension[14]. Studies on the relationship between socio-demographic traits and the anticipated prevalence of hypertension have been conducted using a variety of samples and settings[15]. In order to investigate the risk factors for hypertension in Bangladesh, this study was conducted using a nationally representative sample population. In this study, we looked at the prevalence, consciousness, and management of hypertension as well as the socio-demographic variables that affect these results among adult Bangladeshis.

To explore the spatial characteristics of hypertension clusters, spatial clustering of hypertension prevalence has, however, only occasionally been used, especially at the national level. The objective of our study was to detect any districts that had a particularly high relative risk of hypertension prevalence. The findings will be used by academics, researchers, decision-makers, and members of the public health community to generate new regional plans for hypertension prevention and control.

## Methods

### Data source

Through secondary evaluation of the BDHS 2017–18 data, we were able to do a cross-sectional analysis.[9]. This is the eighth major national survey that provides data on the nation’s demographic and health trends. The list of enumerated areas (EAs) from the 2011 Population and Housing Census of the People’s Republic of Bangladesh, supplied by the Bangladesh Bureau of Statistics (BBS), serves as the survey’s sampling frame (BBS 2011). An EA containing an average size of around 120 residences serves as the survey’s primary sampling unit (PSU). A two-stage, stratified sample of households was used as the foundation for the survey. With a probability corresponding to EA size, 675 EAs were selected in the first phase (250 in communities and 425 in rural communities). The survey was successfully completed in 672 clusters after three clusters (one urban and two rural) were excluded because they had been entirely destroyed by floodwater. These clusters were in Rangpur (one rural cluster), Rajshahi (one rural cluster), and Dhaka (one urban cluster). The survey’s sample size was 20,160 households in total. In the subsample of a quarter of the households, blood samples for glucose testing and blood pressure (BP) measurements were drawn from all men and women age 18 and older (independent of marital status). This subsample contained 12,926 in all (5,583 men and 7,343 women).

### Outcome variables

Our outcomes of interest included the prevalence, knowledge, medication, and control of hypertension. According to the survey, diastolic and systolic blood pressure were observed in millimeters of mercury (mmHg). The use of digital oscillometric BP measurement devices that automatically inflate and deflate the upper arm was completed. The mean of the second and third measures, taken over three measurements spaced at least five minutes apart, served as the individual blood pressure reading. At the time of the survey, a person was classified as having hypertension if their average systolic blood pressure (SBP) was 140 mmHg, their average diastolic blood pressure (DBP) was 90 mmHg or higher, or if they were actively taking antihypertensive medication. The term “awareness of hypertension” was used to describe hypertensive responders who knew they were hypertensive because a doctor or other health professional had informed them of their condition. Self-reported usage of prescription antihypertensive medications by those who have been diagnosed as having high blood pressure is considered to constitute the medication of hypertension. An average SBP and DBP of less than 140 mmHg and 90 mmHg, respectively, among individuals taking antihypertensive medication were considered to be under control of hypertension.

### Explanatory variables

We examined the following as likely variables: place, division of residence, level of education, wealth quintile, gender, age, overweight/obesity, type 2 diabetes, and published information and reports. Age groups for men and women were 18–34, 35–44, 45–54, 55–64, and 65 or older. Gender was classified as male or female. According to the WHO expert consultation, the Asian cut-off point of 18.5 for underweight, 18.5-23.0 for normal weight, 23.0-27.5 for overweight, and 27.5 for obesity was used to classify BMI[16] because Asians are more likely than Caucasians to develop type 2 diabetes as well as cardiovascular disease with BMIs below the WHO’s current standard. Insulin-lowering medication, self-reported use, or a fasting blood sugar level above or equivalent to 7.0 mmol/L were regarded as diabetes indications. There were four categories for education level: no formal education, primary, secondary, and college or higher. The wealth quintile was classified as poorest, poorer, middle, richer, and richest. Places of residence were categorized as urban and rural. The eight residential divisions were Barishal, Chittagong, Rajshahi, Rangpur, Dhaka, Khulna, Mymensingh, and Sylhet.

### Statistical analysis

For the socio-demographic data and additional potential predictors, a descriptive analysis was conducted. We calculated the presence of hypertension, consciousness, medical treatment, and control. The imprecise estimates were adjusted with a sample weight and an effective survey design. The confidence interval (CI) of 95% was employed to represent the result, and the data was considered statistically significant if p< 0.05. Stata (StataCorp, 2013) software was used for all descriptive and statistical data analysis (version 13.1).

### Spatial analysis

To measure the spatial proximity between each potential pair of locations, the spatial weight matrix (w) has been developed. Various methods, including the Queen’s, Rook’s, and Bishop’s methods, can be used to calculate the matrix depending on how the neighbors are defined. We calculated the matrix here using the Queen’s method[17]. This weight matrix may be interpreted as a distance-based weight or as a measure of contiguity between cells. We conducted a distance-based weight in this study[18]. As a standard measure of spatial autocorrelation, we first calculated global and local Moran’s I. The Moran’s I value are in the range of −1 to 1[19]. When identical values cluster together, the value “1” denotes perfect positive spatial autocorrelation, but when dissimilar values cluster together, the value “-1” denotes perfect negative spatial autocorrelation. Also, it is implied that there are random spatial feature distributions if the local Moran’s I value is equal to or close to 0. Additionally, the local Moran’s index, also known as the local indicator of spatial association (LISA)[20], which measures the spatial autocorrelation of an event at a specific location, was calculated. Moreover, the regional Moran’s is utilized to determine the geographic distribution of hypertension clusters and outliers using five fundamental classifications:

1. High-High: The High-High classification refers to districts with a high prevalence of hypertension that are flanked by other counties with a high prevalence. It is also known as a “hot spot.”
2. Low-Low: The Low-Low classification corresponds to districts displaying low levels of hypertension prevalence that are surrounded by counties with similarly low values. It’s called “cold spots.”
3. High-Low: According to the High-Low (spatial outliers) classification, districts with high rates of hypertension prevalence are surrounded by those with low rates.
4. Low-High: The Low-High (spatial outliers) classification refers to districts with a low prevalence of hypertension that are surrounded by a high prevalence.
5. Not-significant: districts that have spatial patterns but are not statistically significant.

With a 95% confidence interval (a 5% level of significance), we plotted the Lisa significance map[21]. The statistical level at which each region can be considered to have made a significant contribution to the results of the global spatial autocorrelation is shown on this map. All analyses were done using R (version 4.2.1).

## Results

The analysis had a total of 12,926 participants, of whom 3551 were hypertensive. Table 1 shows that the participants were 39.7 years old on average, with a standard error of 0.16. 58.8% of all hypertensive people are female, and 41.2% are male. The average BP was 80.2 mmHg at the diastolic level and 122.4 mmHg at the systolic level (Table 1). 27.4% (n = 3542) of total participants are from urban areas, and 72.6% (n = 9384) of total participants are from rural areas. Around 26.6% (n = 3444) of the participants lacked a formal education.

**Table 1.**
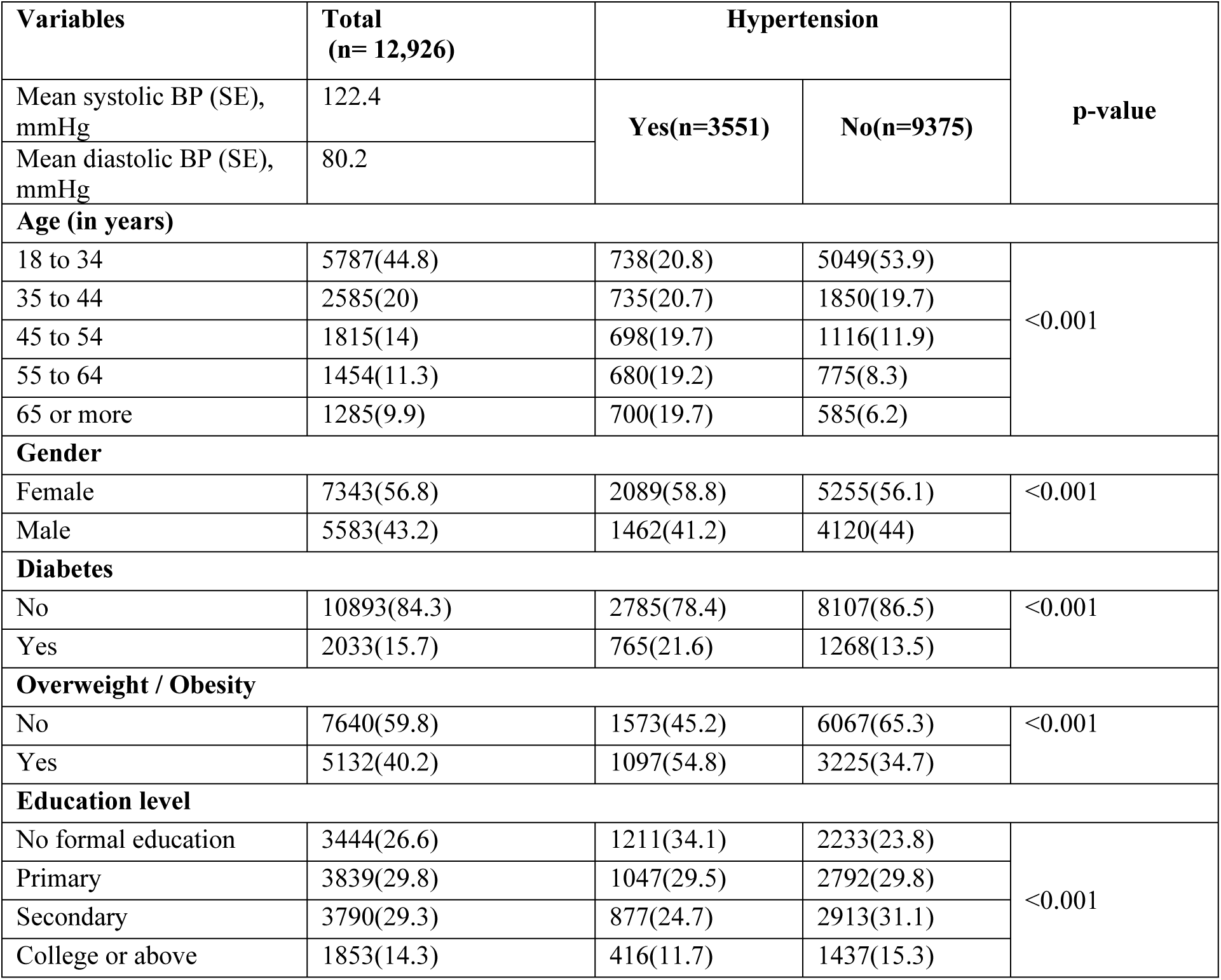
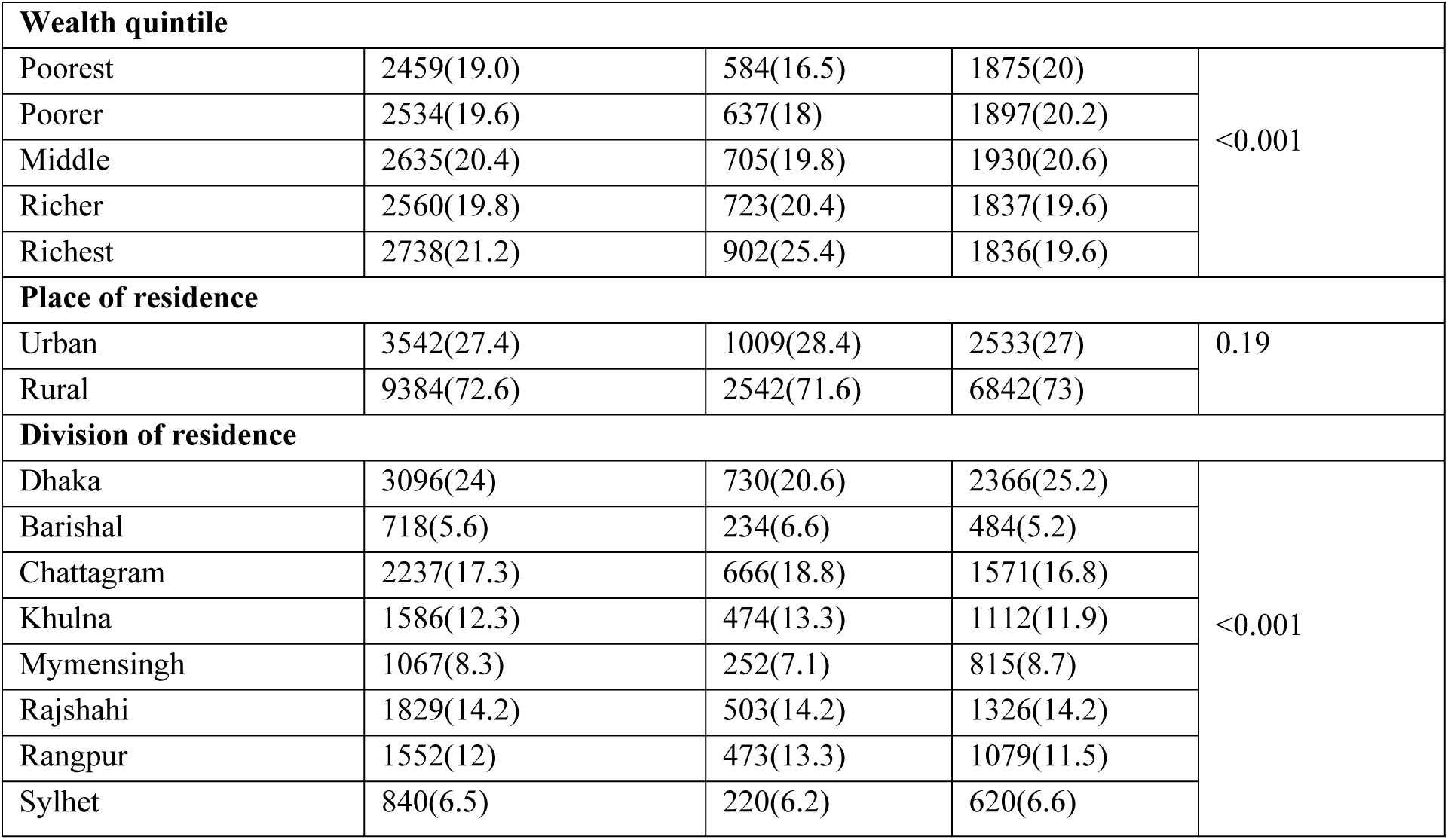
Distribution of the study sample according to presence of hypertension.

According to background characteristics, Table 2 displays the prevalence, consciousness, medication and control of hypertension by gender. Among men, the older age group (65 and older) has a greater prevalence of hypertension, which is close to 50.64% (95% CI: 46.67%–54.61%). The age group from 18 to 34, which is the youngest age group, has the lowest prevalence of hypertension (13.24%, 95% CI: 11.64%–15.03%). And for female participants, compared to younger age groups, older age groups had a prevalence of hypertension that was nearly 59.15% (95% CI: 54.66%– 63.49%). And among the youngest age groups, those between 18 and 34, there was the lowest rate of hypertension (12.46%, 95% CI: 11.32%–13.70%). The overall hypertension prevalence was higher in urban areas for both male (27.93%, 95% CI: 25.45%–30.56%) and female (28.91%, 95% CI: 26.99%–30.91%) participants. The analysis shows that obese individuals, both males (41.58%, 95% CI: 36.09%–47.29%) and females (44.50%, 95% CI: 41.56%– 47.48%), have an increased risk of developing hypertension.

**Table 2:**
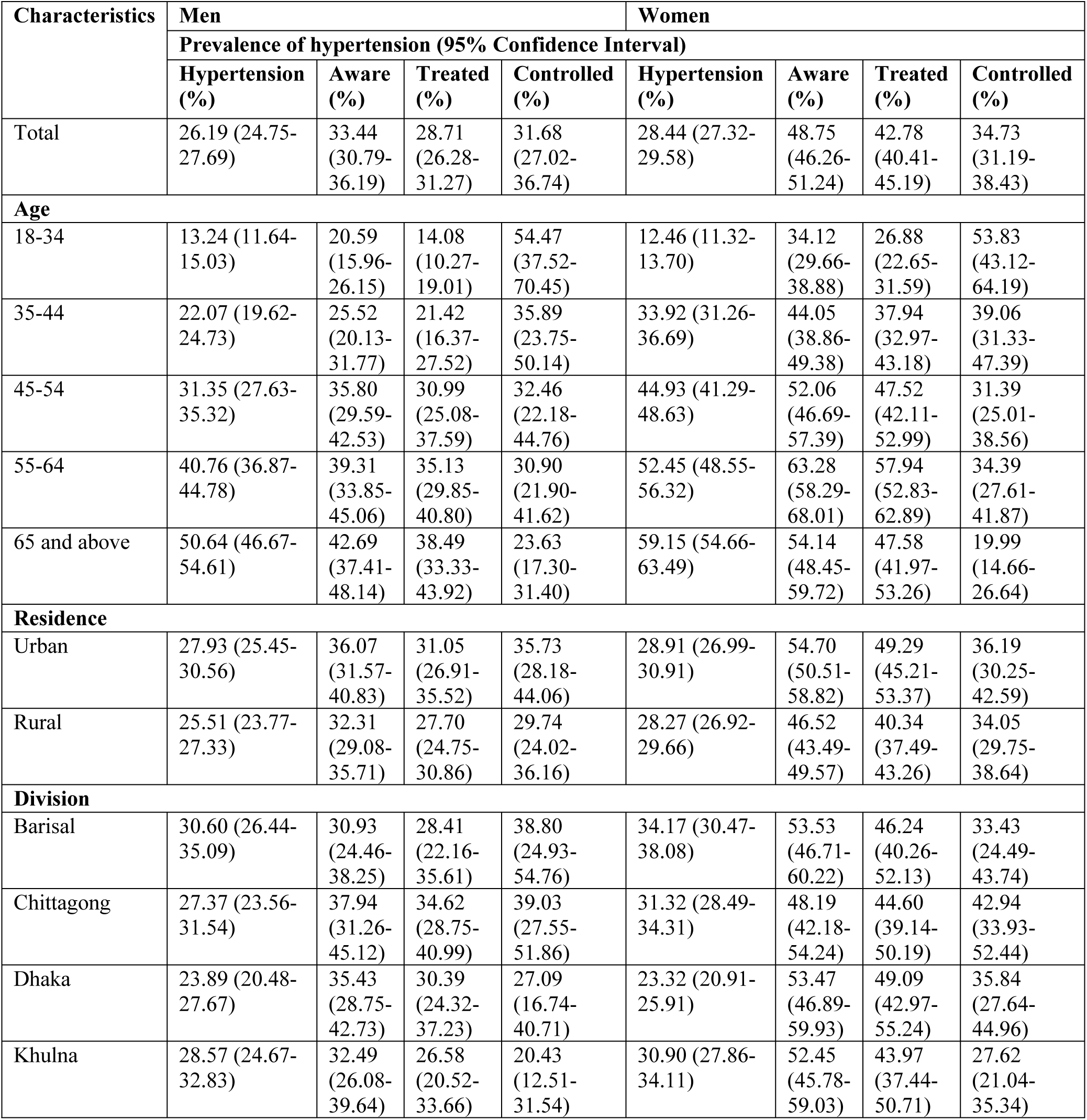
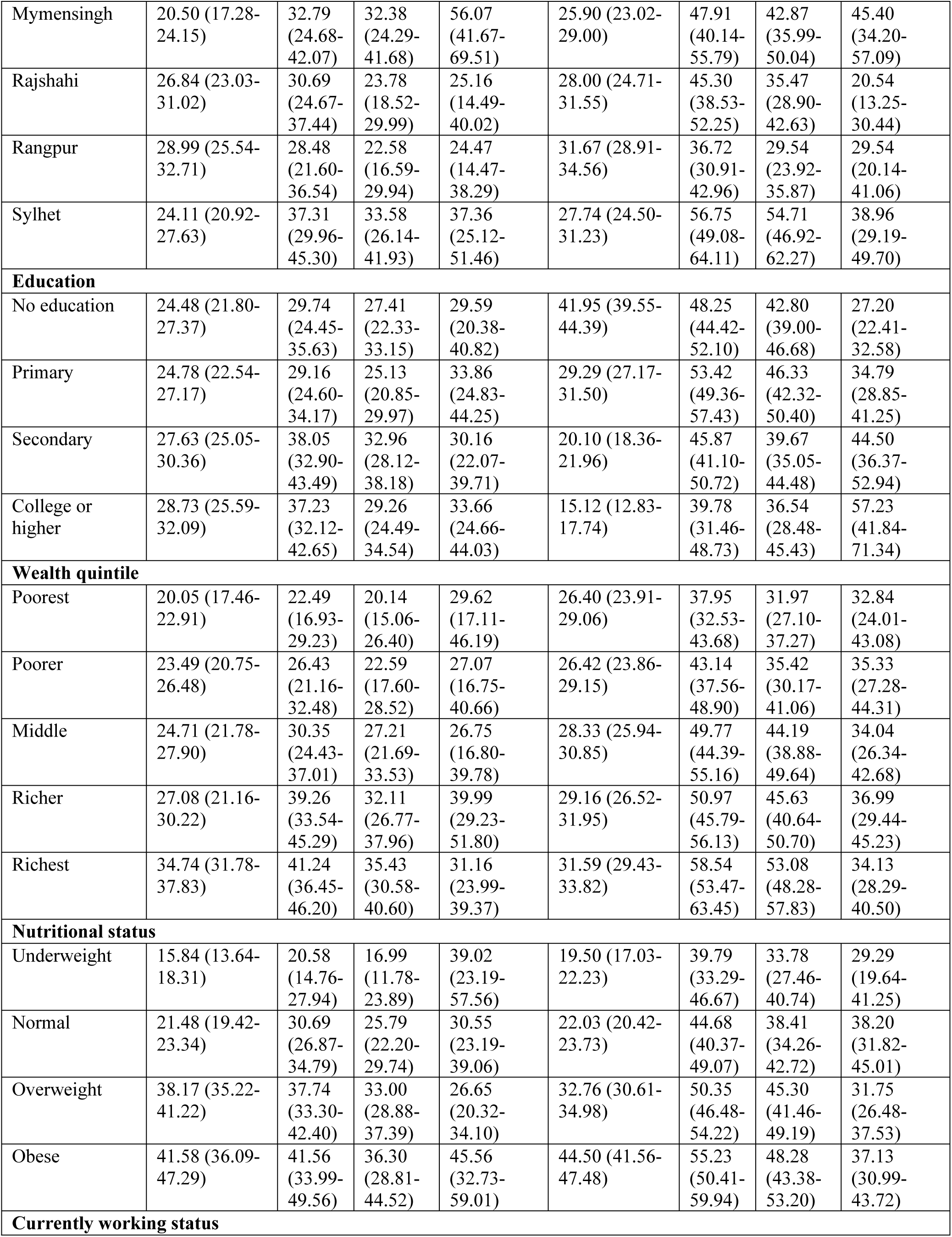
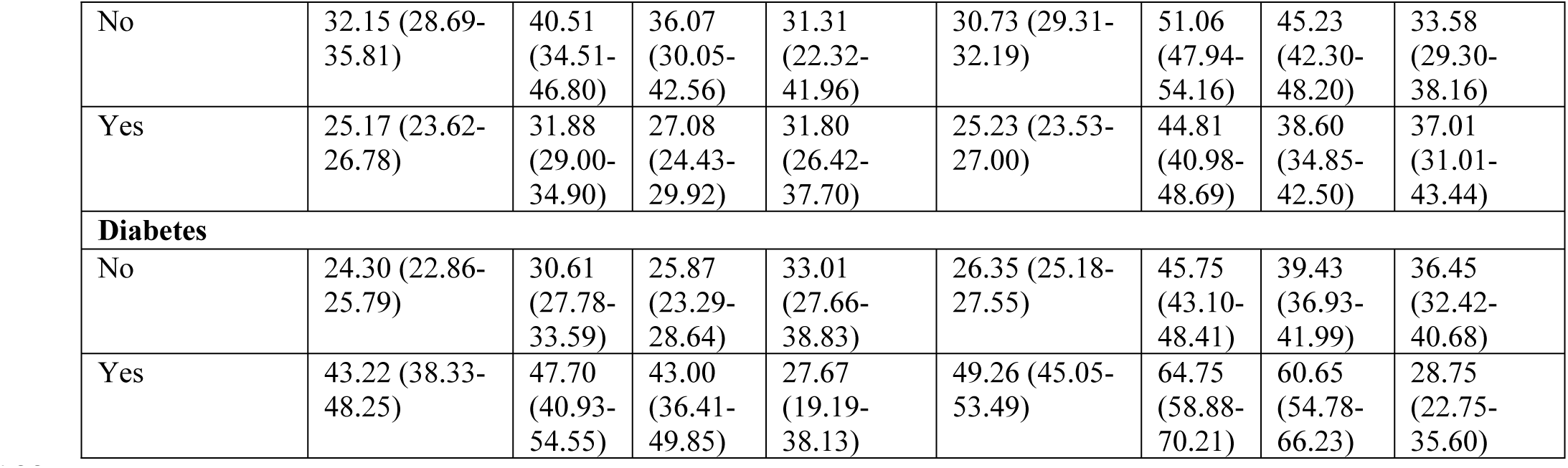
The prevalence, awareness, and control of hypertension by gender.

In all age categories, among the overall population of hypertensives, more women than men are aware of their condition and receive treatment for it. However, only in the age categories of 35–44 and 55–64 do women have a higher prevalence of hypertension control than do men. Once more, urban areas have greater rates of hypertension awareness (Men: 36.07%, 95% CI: 31.57%-40.83%; Women: 54.70%, 95% CI: 50.57%-58.82%), treatment (Men: 31.05%, 95% CI: 26.91%-35.52%; Women: 49.29%, 95% CI: 45.21%-53.37%), and control (Men: 35.73%, 95% CI: 28.18%-44.06%; Women: 36.19%, 95% CI: 30.25%-42.59%) than rural ones. In addition, women are probable to have more hypertension consciousness, medication, and control than men in both rural and urban regions. The lowest levels of awareness, medication, and control of hypertension are found among the poorest individuals of both sexes. According to the study, both men and women who are obese are more probable to be conscious of high BP, receive medical treatment, and have control over it. Table 2 displays all of the findings.

Fig 1 and Fig 2 show how men and women in Bangladesh are divided in terms of their awareness of hypertension and their treatment status. The four levels of the pie chart—“Aware, not treated,” “Aware, controlled,” “Aware, treated,” and “Unaware”—are present in each of the eight divisions. In every district, the pie chart indicates that the majority of men and women with hypertension are unaware that they have the medical condition. The number of people with hypertension in a district is indicated by the size of the pie chart. There are more hypertensive individuals in the area where the pie chart is larger.

**Fig 1:**
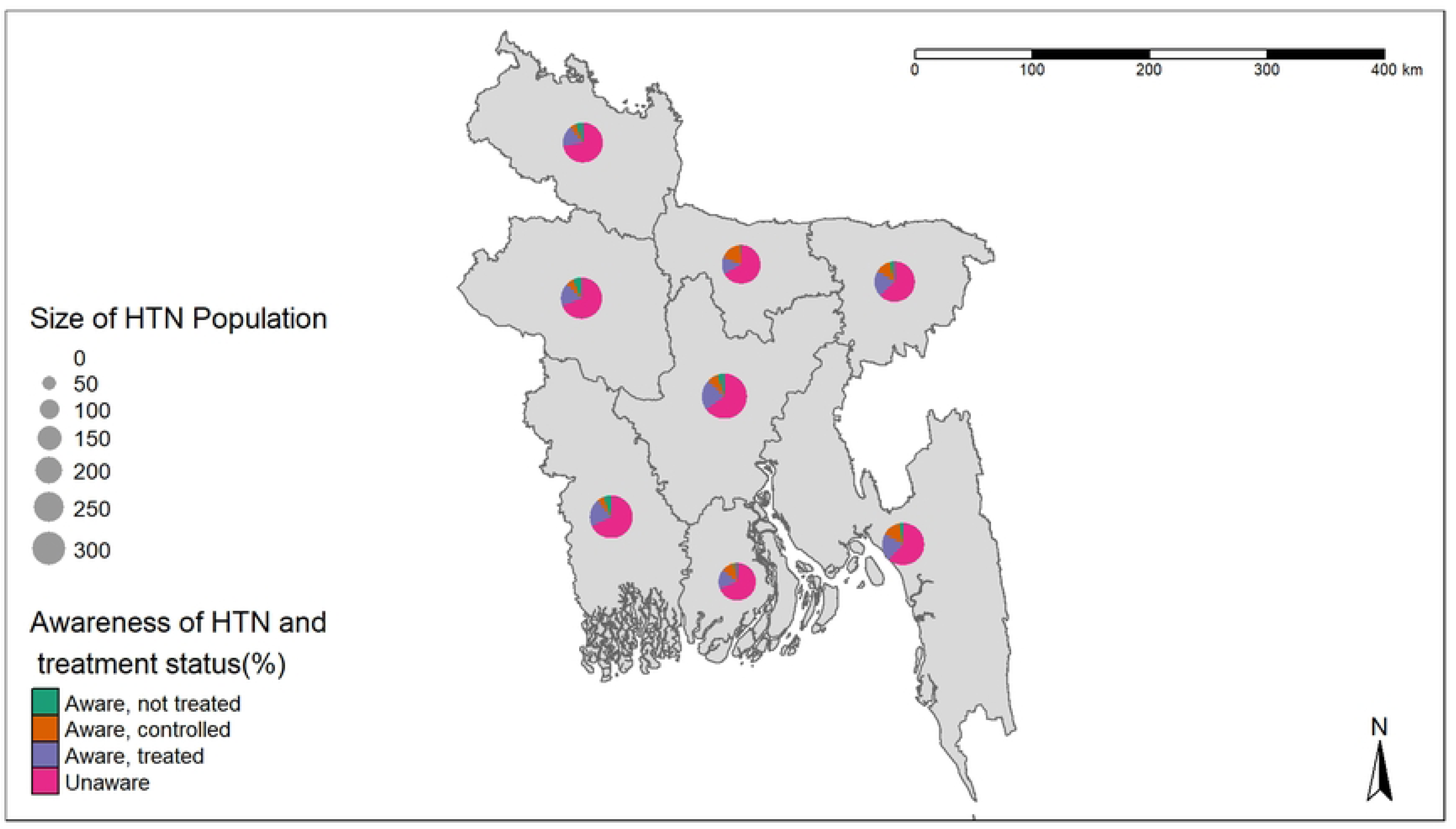
Awareness of hypertension and treatment status for men in Bangladesh.

**Fig 2:**
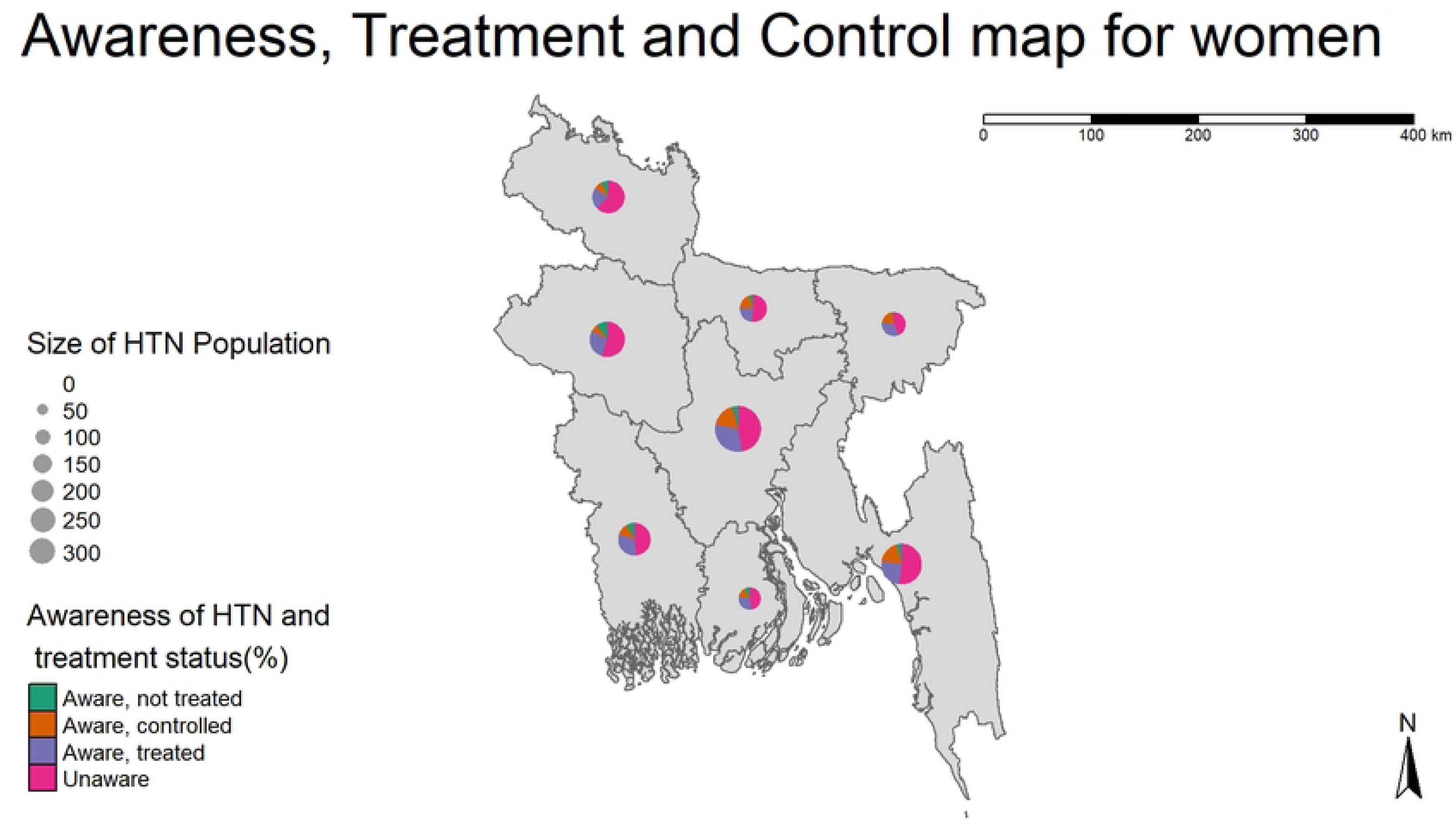
Awareness of hypertension and treatment status for women in Bangladesh.

Local Moran’s I and univariate clustering using the local indicator of spatial association (LISA), as well as significant maps of the prevalence of hypertension, were created to study the spatial dependency and clustering of hypertension in Bangladesh. For the dataset, Moran’s I is 0.272, which denotes a spatial autocorrelation that is positive. Positive Moran’s I point out that the values in the dataset have a tendency to spatially cluster, which means that high values tend to cluster close to other high values and low values tend to cluster close to other low values. The univariate LISA cluster map (Fig 3) and LISA significance map (Fig 4) demonstrate that the hotspot points (4 districts) are located in the districts of Bagerhat, Munshiganj, Pirojpur, and Rajbari; on the other hand, the districts with cold spots (8 districts) are Bandarban, Habiganj, Kishoreganj, Mymensingh, Netrokona, Sherpur, Sunamganj, and Sylhet. The statistical level at which any region can be considered to be significantly contributing to the results of the global spatial autocorrelation is displayed on the LISA significance map (Fig 4). Statistical significance is determined at the 0.001 (or 0.01 or 0.05) level if an actual LISA score is in the top 0.1% (or 1% or 5%) of scores connected to that area under randomization. An outcome that is statistically relevant can be either extremely high or extremely low.

**Fig 3:**
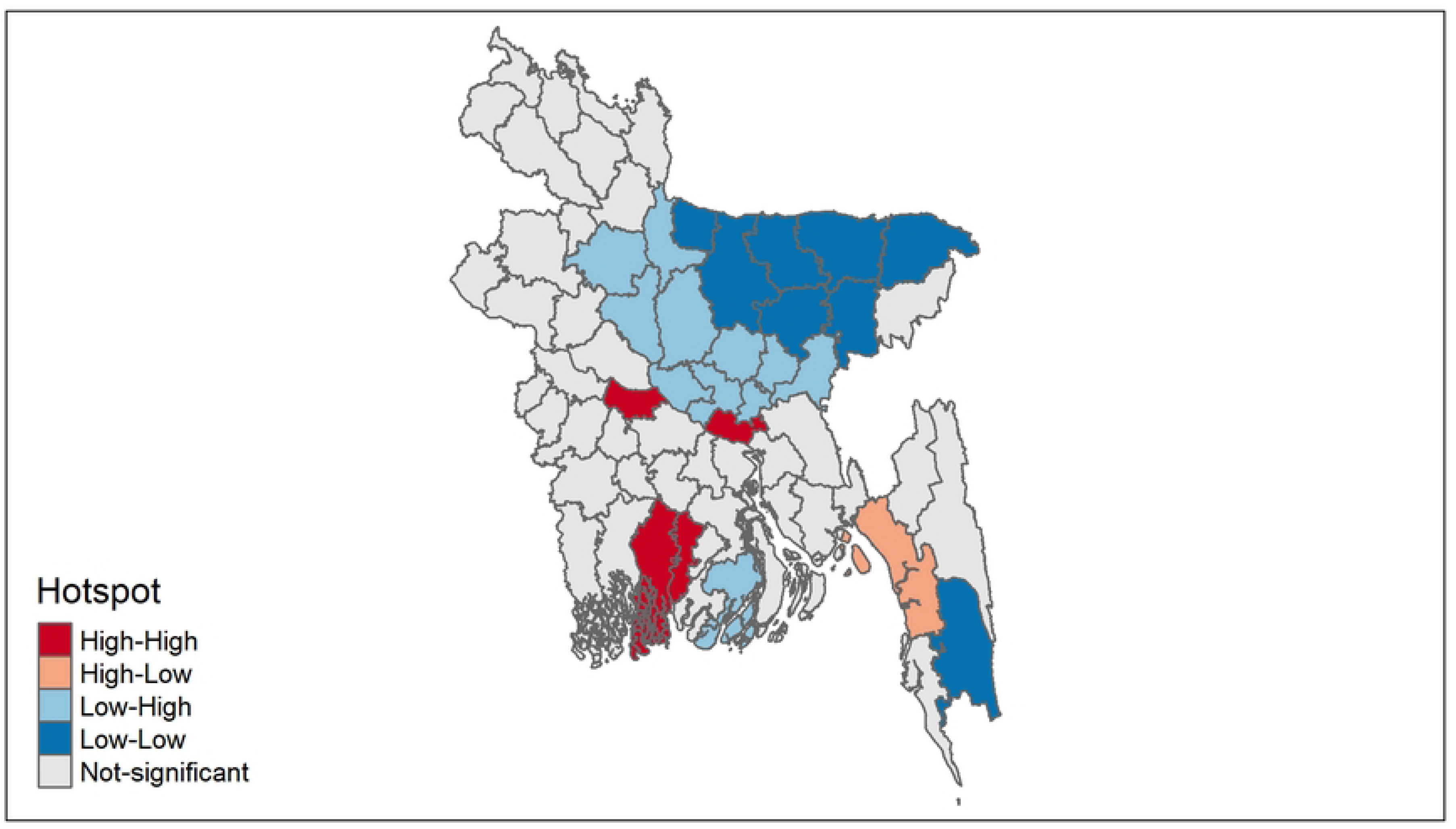
LISA Cluster Map.

**Fig 4:**
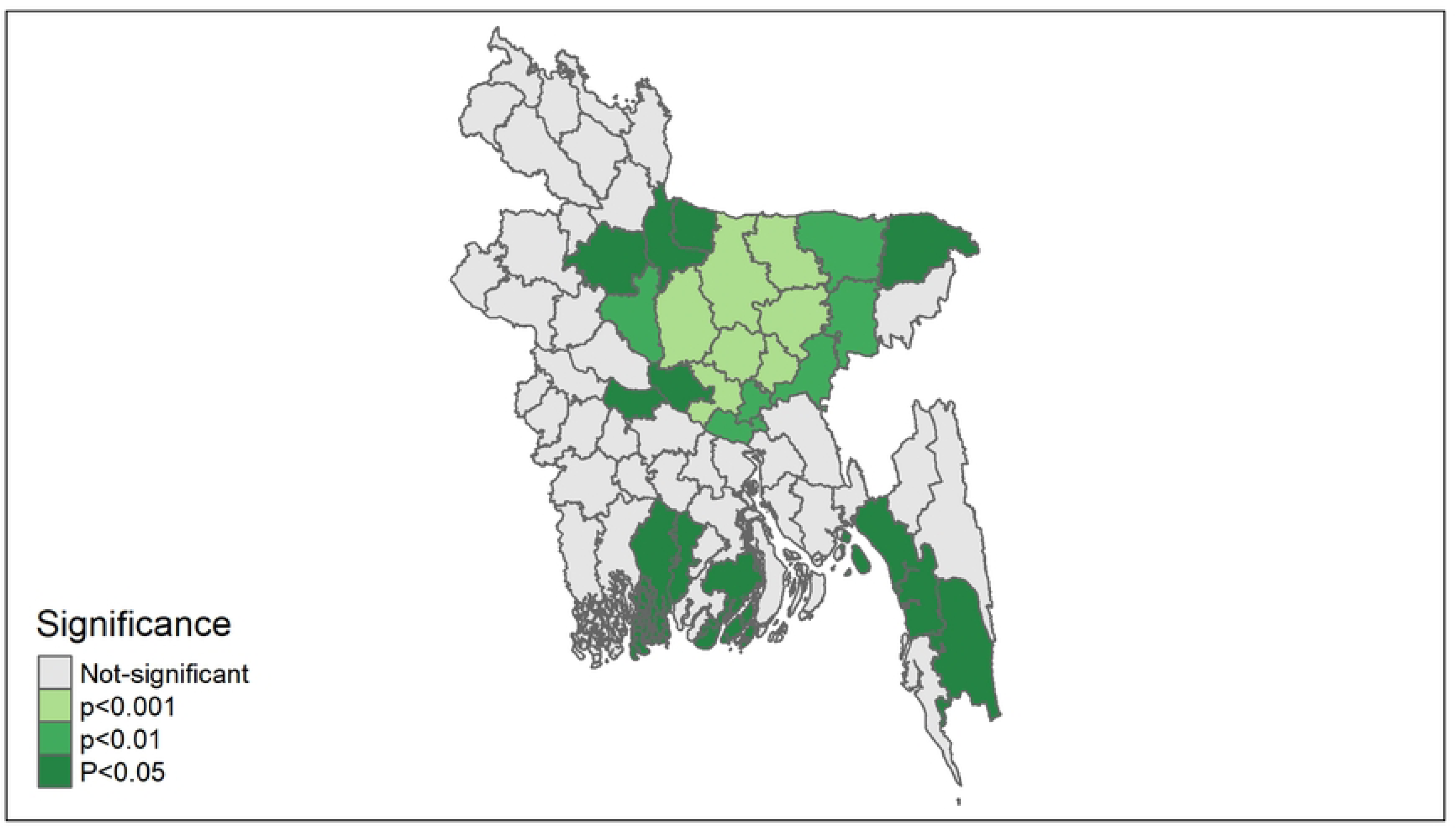
LISA Significance Map.

## Discussion

Using BDHS 2017–18 data, we calculated the prevalence of the condition, along with its consciousness, medication, and control of the condition, and identified risk factors related to it. According to our data, Bangladesh has a significant prevalence of hypertension. More than one in four adults in Bangladesh might suffer from hypertension, according to this study. In contrast, one in every three adults in low- and middle-income countries (LMICs) suffers from hypertension[22,23]. We also discovered other related factors for these outcomes, including age, gender, living in different locations and divisions, education, wealth quintiles, diabetes, and being overweight or obese.

Among total hypertensive individuals, 33.44% of men (95% CI: 30.79%–36.19%) and 48.75% of women (95% CI: 46.26%–51.24%) were conscious of their condition, indicating that women were more conscious of hypertension than men. In men, 28.71% (95% CI: 26.28%–31.27%) of those who had hypertension were being treated, and 31.68% (95% CI: 27.02%–36.74%) had the condition under control. In contrast, among women, 34.73% (95% CI: 31.19%– 38.43%) of hypertensive patients had their hypertension under control, while 42.78% (95% CI: 40.41%–45.19%) were receiving treatment for their condition. We can conclude from the data that women were more likely than men to undergo treatment for their hypertension and keep control over it.

We also observed comparable results when comparing our findings with those of reports from other neighboring areas. The overall prevalence of hypertension in India was 47.5% in males and 48.4% in females[24]. In Sri Lanka, 28.1% (95% CI: 25.9%–30.3%) of men and 28.2% (95% CI: 26.4%–30.0%) of women were hypertensive[25]. 29.7% (95% CI: 24.1%–35.4%) of men and 31.5% (95% CI: 27.5%–38.4%) of women in the Maldives had hypertension prevalence[26]. The overall prevalence of hypertension in Pakistan was 24.99% in males and 24.76% in females[27]. And in China, 30.6% of males and 35.5% of females had hypertension prevalence[28]. In the Indonesian study population, the overall prevalence of hypertension was reported at 33.4% (95% CI: 32.7%–34.0%), 31.0% (95% CI: 30.2%–31.9%) for men, and 35.4% (95% CI: 34.6%–36.3%) for women.[29].

As in Bangladesh, the prevalence of hypertension has increased in these countries as well[30,31]. In the majority of countries, women were more likely than men to have hypertension[24,26,27,29,30,32]. Lifestyle, environment, and food habits differ significantly between urban and rural areas in developing countries like Bangladesh[33]. Rapidly expanding unplanned urbanization, as witnessed in emerging nations, has the potential to cause changes in lifestyles (such as changes in stress levels, physical activity, food, and environment)[34]. In our study, a greater pooled prevalence of hypertension was found among the urban population, demonstrating the strong relationship between these factors and a higher prevalence of hypertension.

Hypertension was associated with gender, age, BMI, type 2 diabetes, and several administrative areas in the country. According to our study, obesity is highly associated with hypertension[35,36].

If we compare awareness of hypertension with that of other Asian countries, 43.79% of men and 27.41% of women are aware of being hypertensive[37]. That indicates men are more aware than women in the Maldives, which is the opposite scenario compared to Bangladesh. In Pakistan, awareness and control of hypertension among women (Awareness: 67.8%; Control: 24.3%) are higher than among men (Awareness: 58%; Control: 20.1%)[38]. Also in China, women (51.0%) are more aware of hypertension than men (35.1%)[39]. Treatment (Men: 26.8%; Women: 42.3%) and control (Men: 7.6%; Women: 11.3%) are also higher among women than among men in China[39]. Also in Indonesia, women are more inclined to be conscious of their condition, receive treatment, and have control over hypertension than men[40].

Numerous factors, including a nation’s economic situation[41], age[42], wealth quintile[43], obesity[44], diabetes[45], and education level[46], had an extensive impact on the occurrence, knowledge, treatment, and control of hypertension. People with diabetes or obesity may require more medical attention than those without these conditions, which may account for their greater concern about the problem[47]. People in rural regions and those with lower levels of wealth have generally lower levels of awareness; this difference in awareness may be due to variations in access to, availability of, and behavior with regard to health care[9].

Researchers have recently used statistical methods and spatial analysis to investigate the spatial variation in hypertensive conditions. For this spatial analysis, local and global Moran’s I were obtained. The Geographical Autocorrelation (the global Moran’s I) simultaneously computes the spatial autocorrelation on the basis of feature positions and values [48]. It examines if the pattern is random, dispersed, or clustered when a set of qualities and corresponding attributes are present[49]. There is no difference between the local and global Moran’s I. Local Moran’s I also discovers geographical clustering of the values in addition to outliers of the data[20,50].

Instead of directly comparing the attribute values of neighboring features to each other, Moran’s index compares the values for every characteristic in the pair to the average value for the dataset. The values of the features are clustered if the average difference between neighboring values is less than the mean between all characteristics[51]. The Moran’s index result showed that the spatial autocorrelation was positive for the prevalence of hypertension in Bangladesh. Similar values for this data have a tendency to cluster together in response to positive spatial autocorrelation[17,52].

Moran’s I statistic was used to perform a hotspot analysis as well. According to our data, the hotspot analysis shows the high-high (hotspot) and low-low (cold spot) districts based on the prevalence of hypertension[53]. The cluster points were displayed on a LISA cluster diagram that was created via local Moran’s I.[54]. The LISA cluster map displays spatial outliers (high-low and low-high areas) as well as statistically not significant areas in addition to hot spots and cold spots. Four districts are shown on the LISA cluster map as hotspots, indicating that there is a high prevalence of hypertension in those four districts. On the other hand, the map has eight districts as cold spots, indicating those eight districts have a low prevalence of hypertension.

Locations with noticeable local statistics were pictured on the map of local significance in order of their level of significance. Significance was based on a permutation test. It displays the districts that have a significant impact on spatial autocorrelation[17,21,55]. The value becomes significant at a level of 0.1%, 1%, or 5%.

There are many benefits to this study. It used a large, regionally representative sample and included all individuals residing in non-institutional housing units across the nation[9]. This study is further strengthened by the utilization of precise measuring techniques for clinical variables such as blood pressure, weight of the person, and height. Additionally, rather than relying on self-reporting, the measurements of hypertension were taken by skilled and experienced health professionals using WHO-recommended techniques. In contrast to other cross-sectional studies conducted in Bangladesh, DHS uses reliable and accepted data collection methods, which reduce measurement error and bias. And 99% of respondents responded in total[9]. Additionally, the decision-making process is enhanced by the spatial analysis. Spatial analysis is used to determine where and how common hypertension is under specific circumstances.

Even though our study had many positive aspects, it also had some shortcomings. Our capacity to establish causal relationships is restricted because this study was cross-sectional. Other limitations included the fact that antihypertensive therapy was self-reported and that our definitions of hypertension and blood pressure management were established on blood pressure data from one point in time. Furthermore, a sizeable portion of those who had hypertension were not questioned about their medication status since they were not aware that they had hypertension. Despite the fact that diet, exercise, and smoking are significant predictors of hypertension, these data were not gathered since they could not be taken into consideration in the analyses. The smaller geographic units should be the focus of future data collection activities. The boundaries of the subdistricts, which reflect administrative requirements instead of the actual spatial distribution, are another downside of our approach. Due to the fact that computations are typically based on the spatial neighborhood surrounding each feature, some spatial statistical approaches may call for information on characteristics that refer to spatial units outside the study area. If certain boundary data are not obtainable, this problem reflects a type of data incompleteness. The Getis-Ord General G-statistic was not utilized in this analysis to determine the concentration of values, which was another limitation.

## Conclusion

Although hypertension is fairly common in Bangladesh, knowledge, treatment, and management remain comparatively low. Therefore, efforts to raise awareness about hypertension, medication, and control need to be given high importance in order to reduce the overall hypertension prevalence and improve hypertension control in Bangladesh, regardless of the risks connected with it and its management. In spatial pattern analysis, the LISA statistic can be used to investigate the prevalence of hypertension in relation to clusters. Additionally, LISA can identify the central focus of hypertension clusters with high and low prevalence. In terms of constructive strategies and actions with regard to the hotspot areas, the health and human development sectors must be given priority. These should include health promotion and hypertension prevention, proactive case reporting, rapid treatment and rehabilitation for those suffering from hypertension and its complications, and preventative case reporting. The map could be used as a reference to identify risk factors in the various locations, which could lead to more effective hypertension prevention and control initiatives that are applicable to the socio-economic context.

## Data Availability

The data used in this study can be found by requesting to the Demographic and Health Survey. The dataset can be found at https://dhsprogram.com/data/.

## Acknowledgement

For permission to use the BDHS 17_18 dataset, the authors expressed their gratitude to MEASURE DHS.

## Reference

[1] GBD 2017 Risk Factor Collaborators. Global, regional, and national comparative risk assessment of 84 behavioural, environmental and occupational, and metabolic risks or clusters of risks for 195 countries and territories, 1990-2017: a systematic analysis for the Global Burden of Disease Study 2017. Lancet Lond Engl 2018;392:1923–94. 10.1016/S0140-6736(18)32225-6.

[2] Thomas H, Diamond J, Vieco A, Chaudhuri S, Shinnar E, Cromer S, et al. Global Atlas of Cardiovascular Disease 2000-2016: The Path to Prevention and Control. Glob Heart 2018;13:143–63. 10.1016/j.gheart.2018.09.511.

[3] Kearney PM, Whelton M, Reynolds K, Muntner P, Whelton PK, He J. Global burden of hypertension: analysis of worldwide data. Lancet Lond Engl 2005;365:217–23. 10.1016/S0140-6736(05)17741-1.

[4] Lawes CMM, Vander Hoorn S, Law MR, Elliott P, MacMahon S, Rodgers A. Blood pressure and the global burden of disease 2000. Part II: estimates of attributable burden. J Hypertens 2006;24:423–30. 10.1097/01.hjh.0000209973.67746.f0.

[5] Zhou B, Bentham J, Cesare MD, Bixby H, Danaei G, Cowan MJ, et al. Worldwide trends in blood pressure from 1975 to 2015: a pooled analysis of 1479 population-based measurement studies with 19·1 million participants. The Lancet 2017;389:37–55. 10.1016/S0140-6736(16)31919-5.

[6] Mills KT, Bundy JD, Kelly TN, Reed JE, Kearney PM, Reynolds K, et al. Global Disparities of Hypertension Prevalence and Control: A Systematic Analysis of Population-Based Studies From 90 Countries. Circulation 2016;134:441–50. 10.1161/CIRCULATIONAHA.115.018912.

[7] Chowdhury MZI, Rahman M, Akter T, Akhter T, Ahmed A, Shovon MA, et al. Hypertension prevalence and its trend in Bangladesh: evidence from a systematic review and meta-analysis. Clin Hypertens 2020;26:10. 10.1186/s40885-020-00143-1.

[8] Al Kibria GM, Swasey K, Hasan MdZ, Choudhury A, Gupta RD, Abariga SA, et al. Determinants of hypertension among adults in Bangladesh as per the Joint National Committee 7 and 2017 American College of Cardiology/American Hypertension Association hypertension guidelines. J Am Soc Hypertens JASH 2018;12:e45–55. 10.1016/j.jash.2018.10.004.

[9] Niport NI of PR and T-, Welfare M of H and F, ICF. Bangladesh Demographic and Health Survey 2017-18 2020.

[10] Malik A. Congenital and acquired heart diseases: (A survey of 7062 persons). Bangladesh Med Res Counc Bull 1976;2:115–9.

[11] Saquib N, Saquib J, Ahmed T, Khanam MA, Cullen MR. Cardiovascular diseases and type 2 diabetes in Bangladesh: a systematic review and meta-analysis of studies between 1995 and 2010. BMC Public Health 2012;12:434. 10.1186/1471-2458-12-434.

[12] Islam AKMM, Majumder A a. S. Coronary artery disease in Bangladesh: a review. Indian Heart J 2013;65:424–35. 10.1016/j.ihj.2013.06.004.

[13] Alam DS, Chowdhury MAH, Siddiquee AT, Ahmed S, Niessen LW. Awareness and control of hypertension in Bangladesh: follow-up of a hypertensive cohort. BMJ Open 2014;4:e004983. 10.1136/bmjopen-2014-004983.

[14] Rahman M, Williams G, Al Mamun A. Gender differences in hypertension awareness, antihypertensive use and blood pressure control in Bangladeshi adults: findings from a national cross-sectional survey. J Health Popul Nutr 2017;36:23. 10.1186/s41043-017-0101-5.

[15] Zhang B, Menzin J, Friedman M, Korn JR, Burge RT. Predicted coronary risk for adults with coronary heart disease and low HDL-C: an analysis from the US National Health and Nutrition Examination Survey. Curr Med Res Opin 2008;24:2711–7. 10.1185/03007990802363198.

[16] Lim JU, Lee JH, Kim JS, Hwang YI, Kim T-H, Lim SY, et al. Comparison of World Health Organization and Asia-Pacific body mass index classifications in COPD patients. Int J Chron Obstruct Pulmon Dis 2017;12:2465–75. 10.2147/COPD.S141295.

[17] Anselin L, Syabri I, Kho Y. GeoDa : An Introduction to Spatial Data Analysis. Geogr Anal 2006;38:5–22. 10.1111/j.0016-7363.2005.00671.x.

[18] Anselin L, Smirnov O. Efficient Algorithms for Constructing Proper Higher Order Spatial Lag Operators*. J Reg Sci 1996;36:67–89. 10.1111/j.1467-9787.1996.tb01101.x.

[19] Moran PAP. Notes on Continuous Stochastic Phenomena. Biometrika 1950;37:17–23. 10.2307/2332142.

[20] Local Spatial Autocorrelation (1) 2022. https://geodacenter.github.io/workbook/6a_local_auto/lab6a.html (accessed November 30, 2022).

[21] Project 4: Local Indicators of Spatial Association | GEOG 586: Geographic Information Analysis 2022. https://www.e-education.psu.edu/geog586/node/673 (accessed November 30, 2022).

[22] Sarki AM, Nduka CU, Stranges S, Kandala N-B, Uthman OA. Prevalence of Hypertension in Low- and Middle- Income Countries: A Systematic Review and Meta-Analysis. Medicine (Baltimore) 2015;94:e1959. 10.1097/MD.0000000000001959.

[23] Mills KT, Bundy JD, Kelly TN, Reed JE, Kearney PM, Reynolds K, et al. Global Disparities of Hypertension Prevalence and Control. Circulation 2016;134:441–50. 10.1161/CIRCULATIONAHA.115.018912.

[24] Gupta PC, Gupta R, Pednekar MS. Hypertension prevalence and blood pressure trends in 88 653 subjects in Mumbai, India. J Hum Hypertens 2004;18:907–10. 10.1038/sj.jhh.1001763.

[25] Rannan-Eliya RP, Wijemunige N, Perera P, Kapuge Y, Gunawardana N, Sigera C, et al. Prevalence and Associations of Hypertension in Sri Lankan Adults: Estimates from the SLHAS 2018–19 Survey Using JNC7 and ACC/AHA 2017 Guidelines. Glob Heart 2022;17:50. 10.5334/gh.1135.

[26] Aboobakur M, Latheef A, Mohamed AJ, Moosa S, Pandey RM, Krishnan A, et al. Surveillance for non-communicable disease risk factors in Maldives: results from the first STEPS survey in Male. Int J Public Health 2010;55:489–96. 10.1007/s00038-009-0114-y.

[27] Shah N, Shah Q, Shah AJ. The burden and high prevalence of hypertension in Pakistani adolescents: a meta-analysis of the published studies. Arch Public Health 2018;76:1–10. 10.1186/s13690-018-0265-5.

[28] Xu L, Lai D, Fang Y. Spatial analysis of gender variation in the prevalence of hypertension among the middle-aged and elderly population in Zhejiang Province, China. BMC Public Health 2016;16:447. 10.1186/s12889-016-3121-y.

[29] Peltzer K, Pengpid S. The Prevalence and Social Determinants of Hypertension among Adults in Indonesia: A Cross-Sectional Population-Based National Survey. Int J Hypertens 2018;2018:5610725. 10.1155/2018/5610725.

[30] World Health Organization. Noncommunicable diseases country profiles 2018. Geneva: World Health Organization; 2018.

[31] Singh RB, Suh IL, Singh VP, Chaithiraphan S, Laothavorn P, Sy RG, et al. Hypertension and stroke in Asia: prevalence, control and strategies in developing countries for prevention. J Hum Hypertens 2000;14:749–63. 10.1038/sj.jhh.1001057.

[32] Malavige GN, de Alwis NMW, Weerasooriya N, Fernando DJS, Siribaddana SH. Increasing diabetes and vascular risk factors in a sub-urban Sri Lankan population. Diabetes Res Clin Pract 2002;57:143–5. 10.1016/s0168-8227(02)00015-3.

[33] Chowdhury MZI, Haque MA, Farhana Z, Anik AM, Chowdhury AH, Haque SM, et al. Prevalence of cardiovascular disease among Bangladeshi adult population: a systematic review and meta-analysis of the studies. Vasc Health Risk Manag 2018;14:165. 10.2147/VHRM.S166111.

[34] Prevalence of metabolic syndrome in Bangladesh: a systematic review and meta-analysis of the studies - PubMed 2022. https://pubmed.ncbi.nlm.nih.gov/29499672/ (accessed November 30, 2022).

[35] Jiang S-Z, Lu W, Zong X-F, Ruan H-Y, Liu Y. Obesity and hypertension. Exp Ther Med 2016;12:2395. 10.3892/etm.2016.3667.

[36] Hall JE, do Carmo JM, da Silva AA, Wang Z, Hall ME. Obesity-induced hypertension: interaction of neurohumoral and renal mechanisms. Circ Res 2015;116:991–1006. 10.1161/CIRCRESAHA.116.305697.

[37] Singh S, Srivastava K, Misra SK, Gupta SC, Kaushal SK. A Cross-sectional study of awareness and treatment seeking behaviour of hypertensive people of rural and urban area of Agra District. Indian J Forensic Community Med n.d.:5.

[38] Shafi ST, Shafi T. A survey of hypertension prevalence, awareness, treatment, and control in health screening camps of rural central Punjab, Pakistan. J Epidemiol Glob Health 2017;7:135–40. 10.1016/j.jegh.2017.01.001.

[39] Wang J, Zhang L, Wang F, Liu L, Wang; H, the China National Survey of Chronic Kidney Disease Working Group. Prevalence, Awareness, Treatment, and Control of Hypertension in China: Results From a National Survey. Am J Hypertens 2014;27:1355–61. 10.1093/ajh/hpu053.

[40] Sujarwoto S, Maharani A. Participation in community-based health care interventions (CBHIs) and its association with hypertension awareness, control and treatment in Indonesia. PLOS ONE 2020;15:e0244333. 10.1371/journal.pone.0244333.

[41] Chow CK, Teo KK, Rangarajan S, Islam S, Gupta R, Avezum A, et al. Prevalence, Awareness, Treatment, and Control of Hypertension in Rural and Urban Communities in High-, Middle-, and Low-Income Countries. JAMA 2013;310:959–68. 10.1001/jama.2013.184182.

[42] Al Kibria GM, Gupta RD, Nayeem J. Prevalence, awareness, and control of hypertension among Bangladeshi adults: an analysis of demographic and health survey 2017–18. Clin Hypertens 2021;27:17. 10.1186/s40885-021-00174-2.

[43] Khan MN, Oldroyd JC, Chowdhury EK, Hossain MB, Rana J, Renzetti S, et al. Prevalence, awareness, treatment, and control of hypertension in Bangladesh: Findings from National Demographic and Health Survey, 2017-2018. J Clin Hypertens Greenwich Conn 2021;23:1830–42. 10.1111/jch.14363.

[44] NCD Risk Factor Collaboration (NCD-RisC). Worldwide trends in body-mass index, underweight, overweight, and obesity from 1975 to 2016: a pooled analysis of 2416 population-based measurement studies in 128·9 million children, adolescents, and adults. Lancet Lond Engl 2017;390:2627–42. 10.1016/S0140-6736(17)32129-3.

[45] Chowdhury MAB, Uddin MJ, Khan HMR, Haque MR. Type 2 diabetes and its correlates among adults in Bangladesh: a population based study. BMC Public Health 2015;15:1070. 10.1186/s12889-015-2413-y.

[46] Williams MV, Baker DW, Parker RM, Nurss JR. Relationship of Functional Health Literacy to Patients’ Knowledge of Their Chronic Disease: A Study of Patients With Hypertension and Diabetes. Arch Intern Med 1998;158:166–72. 10.1001/archinte.158.2.166.

[47] Finley CR, Chan DS, Garrison S, Korownyk C, Kolber MR, Campbell S, et al. What are the most common conditions in primary care? Systematic review. Can Fam Physician Med Fam Can 2018;64:832–40.

[48] Spatial Autocorrelation - an overview | ScienceDirect Topics 2022. https://www.sciencedirect.com/topics/computer-science/spatial-autocorrelation (accessed December 3, 2022).

[49] How Spatial Autocorrelation (Global Moran’s I) works—ArcGIS Pro | Documentation 2022. https://pro.arcgis.com/en/pro-app/latest/tool-reference/spatial-statistics/h-how-spatial-autocorrelation-moran-s-i-spatial-st.htm (accessed December 1, 2022).

[50] Monzur T. Local Moran&#39;s I calculation 2022.

[51] Alam M. Spatial Autocorrelation: Neighbors Affecting Neighbors. Medium 2020. https://towardsdatascience.com/spatial-autocorrelation-neighbors-affecting-neighbors-ed4fab8a4aac (accessed December 1, 2022).

[52] GISGeography. Spatial Autocorrelation and Moran’s I in GIS. GIS Geogr 2014.

53. https://gisgeography.com/spatial-autocorrelation-moran-i-gis/ (accessed December 2, 2022).

[53] clubdebambos. What is Hotspot Analysis? Geospatiality 2016. https://glenbambrick.com/2016/01/21/what-is-hotspot-analysis/ (accessed December 2, 2022).

[54] Anselin L. Local Indicators of Spatial Association—LISA. Geogr Anal 1995;27:93–115. 10.1111/j.1538-4632.1995.tb00338.x.

[55] Anselin L. Local Spatial Autocorrelation Clusters 2016:33.

